# Frequency and Clinical Characteristics of Breakthrough Cases Post COVID-19 Vaccine and Predictive Risk Factors in College Students

**DOI:** 10.1101/2023.01.20.23284814

**Authors:** Manal Khudder Abdulrazaq, Ahmed Abd Al Redha Jebur, Baqer Jaafar Ali Hamdan, Ahmed khalid Ibrahim

**Author notes:** **Corresponding author: Manal Khudder Abdulrazaq**, CABH (Med), FRCP (London), Department of Medicine, College of Medicine, University of Baghdad, Professor, Medical City Campus/ Bab-Al Muadham/ Baghdad, Iraq. **Ethics and Reporting:** We declare that all relevant ethical guidelines have been followed. **Funding statement:** None. **Competing Interest Statement:** No competing interest.

## Abstract

**BACKGROUND:** COVID-19 vaccines help protect against infection, severe illness, hospitalization and death. When someone who is vaccinated with either a primary series or a primary series plus a booster dose gets infected with the virus that causes COVID-19, it is referred to as a “vaccine breakthrough infection.”

**OBJECTIVES:** To assess the frequency and clinical characteristics of breakthrough cases of COVID-19 infection and to study the predictive risk factors.

**SUBJECTS&METHODS:** A cross-sectional study was carried out including 604 undergraduate medical and non-medical students in Iraq from 10^th^ of August to 29^th^ of September 2022. Data was collected via an online specific questionnaire and analysed to estimate the frequency of COVID-19 breakthrough cases post vaccination, and number of doses of vaccine used. The association of different factors including age, gender, grade, body mass index, smoking, and comorbidities was also studied as predictive risk factors. We used the data to formulate tables, figures and perform statistical tests in IBM SPSS Statistics 25.

**RESULTS:** Mean age of study sample was 21.78 year ± 3.26 and 339 (56%) were females. In terms of COVID-19 vaccination data, 97 (16%) have received one dose, 459 (76%) two doses and 48 (8%) three doses. Regarding PCR test, 74 (12%) were positive after the first dose compared to 49 (8%) after the second dose. About the symptoms developed, the most frequent were fever in 372 (61.1%), unusual fatigue in 96 (15.79%), chills in 29 (4.77%) and persistent cough in 26 (4.28%). For most predictive factors, results were statistically insignificant.

**CONCLUSIONS:** In current study; demographic factors showed no statistically significant impact on prevalence of COVID-19 breakthrough cases. Despite this; number of participants who develop symptoms after the second dose of vaccine was high; and having 3 or more symptoms. About half of participants showed symptoms even after being fully vaccinated.

## INTRODUCTION

Coronavirus disease 2019 (COVID-19) is caused by SARS-CoV-2, that was first recognized in Wuhan, China, in December 2019. Epidemiology and virologic studies suggest that transmission mainly occurs from symptomatic people to others by close contact through respiratory droplets, by direct contact with infected persons, or by contact with contaminated objects and surfaces (1-4).

Clinical and virologic studies that have collected repeated biological samples from confirmed patients demonstrate that shedding of SARS-CoV-2 is highest in the upper respiratory tract (URT) (nose and throat) early in the course of the disease (5-7), within the first 3 days from onset of symptoms (8-10).

### Breakthrough Cases

Vaccines against (SARS-CoV-2) have been demonstrated to be highly effective in preventing symptomatic infections with (COVID-19) (11,12). A breakthrough infection (also known as a breakthrough case) is a term used to describe a COVID-19 infection that occurs in a fully vaccinated person. The CDC defines a person with a breakthrough infection as someone who has a positive COVID-19 test 14 or more days after receiving a full course of a COVID-19 vaccine. According to data analyzed by the CDC through April 20, more than 87 million people have been fully vaccinated against COVID-19. Of those individuals, 7157 are reported to have had breakthrough infections. Sixty-four percent of the breakthrough infections have occurred in females, and 46% have occurred in people age 60 and over. Thirty-one percent of the people who experienced breakthrough infections had no symptoms, 7% were hospitalized and less than 1% have died (although some of those deaths were also reported as not related to COVID-19). Although the COVID-19 vaccines are highly effective at preventing symptomatic infection and represent a critical aspect of pandemic control, none of the vaccines is 100% effective at preventing infection (13,14).

At the beginning of the (COVID-19) pandemic, it was speculated that SARS-CoV-2 infection would result in lifelong immunity, and reinfections would be unlikely. However, there have been several documented cases of reinfection with SARS-CoV-2 (15). A cohort study reports reinfection rates among a large north Indian health care workers (n = 4978) with SARS-CoV-2 infection in 15 months (including the second wave, which was closely linked to the delta variant). As the result of this study, 124 cases of reinfection (2.5%) were identified. Another study from India from January 22 to 7 October 2020, reported that out of 1300 individuals, 58 (4.5%) were reinfected (16).

Therefore, waning humoral immunity is increasingly recognized as a significant concern. Accordingly, long-term and durable vaccine-induced antibody protection against infection is now a significant challenge facing scientists (17).

### Pathogenesis& risk factors of post-vaccination infections

As previously mentioned, waning immunity after a de novo infection or vaccination can be the reason that some people get infected or reinfected following COVID-19 vaccines. Moreover, some individuals with diminished capacity to produce protective antibodies, such as immunosuppressed patients, are also susceptible to being infected even after being naturally infected with this virus or receiving both vaccine doses (18-21).

Ineffective antibody production, due to relatively ineffective vaccines, an inadequate number of doses, and the time after the vaccination are also involved in the pathogenesis of post-vaccination infections (22). It is not unusual to get infected in the first 14 days following the first dose of the vaccine since protective immunity cannot build within this period (23).

Being younger was associated with an increased risk of a breakthrough infection, also in the multivariate analysis. A higher rate of breakthrough infections in younger adults has also been observed in other cohorts (24-27), after adjustment for exposure to the SARS-CoV-2 virus through their profession or environment. The risk of a breakthrough infection was significantly reduced for those having been previously infected, and for those having received a booster vaccine. Compared with COVID-19 naïve persons, persons having been previously infected and who were re-infected after vaccination were more likely to be completely asymptomatic (28).

## Objectives

- To assess the frequency of breakthrough cases of COVID-19.
- To study the clinical characteristics of breakthrough cases of COVID-19.
- To study the predictive risk factors for breakthrough cases of COVID-19.

## SUBJECTS AND METHODS

### Study population

The study was conducted on undergraduate students of all grades from nine colleges in Iraq, regardless of factors like age, gender, and place of residency.

### Inclusion criteria

All students from different specialities who showed symptoms and or positive PCR test after taking Covid-19 vaccine were included.

### Operational definitions

Age of student was gathered in years, gender as (male/female), the grade as the class of student during 2022. Vaccine choices included all types of vaccines.

### Data sources

During the period from 10^th^ of August to 29^th^ of September 2021, an online forum-based questionnaire was used with several questions relevant to the researchers to assess the general demographic information including age, gender, grade, BMI, smoking (current, non-smoker, and passive), comorbidities (hypertension, DM, heart disease, bronchial asthma, and chronic renal disease). Frequency of breakthrough cases and types of symptoms developed in spite of positive PCR test for COVID-19 was measured. Severity of cases was expressed for the purpose of this study; as being either asymptomatic or having 1-2, three or more symptoms. BMI was calculated as the ratio of weight (kilograms) divided by height (meters) squared (kg/m^2^). Underweight or normal weight is defined as (15.0≤ BMI< 25.0), overweight is defined as (25.0 ≤ BMI < 30.0), obesity is defined as (30.0 ≤ BMI ≤ 40.0), and severe obesity is defined as (BMI>40.0). Data about COVID-19-like symptoms, COVID-19 tests, COVID-19 vaccination status (one or two or three doses) was also collected and analysed.

### Study design

This research composes an online descriptive cross-sectional survey about Covid-19 Breakthrough Cases after taking vaccine doses regardless of doses and type of vaccine.

### Sampling procedure

Convenience sampling.

### Sample size

604 students.

### Data Collection

Data was collected via an online questionnaire using google form was conducted on social media applications (Facebook, Instagram and Telegram).

### Analysis plan

Data was summarized into tables and figures in IBM SPSS Statistics 25. Chi-square was selected as the preferred method of statistical analysis (as it is suitable when dealing with categorical data) with a P-value of 0.05 as the cut-off point for statistical significance.

### Ethical approval

All participants were informed before submission that their answers to the questionnaire will be used for research purposes. This study was approved by the department of Family and Community Medicine in Baghdad College of Medicine.

## RESULTS

### Demographic characteristics of study sample

Participants were classified into three groups according to number of vaccine doses they have received. Mean age was 21.78 year ± 3.26, females were more than males. The majority were non-smokers in 461(76%).

According to the BMI, 399 (66%) were either underweight or normal and only 8 (1%) with severe obesity. The most frequent chronic disease was bronchial asthma found in 48 (8%) followed by systemic hypertension, DM, heart disease and chronic kidney disease in 10 (2%), as shown in Table 1.

**Table 1:** Demographic characteristics of study sample distributed according to number of received doses of COVID-19 vaccine.

| Parameters |  | All Participants<br>N= 604 |  | One does<br>N= 97<br>(16%) |  | Two doses<br>N= 459<br>(76%) |  | Three doses<br>N=48 (8%) |  |
| --- | --- | --- | --- | --- | --- | --- | --- | --- | --- |
| Age/year (Mean ± SD) | | 21.78 ± 3.26 | | 21.46 ± 3.03 | | 21.9 ± 3.4 | | 21.1 ± 1.7 | |
| Smoking | Current Smoker | 90 | (15%) | 16 | 16% | 65 | 14% | 9 | 19% |
|  | Non smoker | 461 | (76%) | 76 | 78% | 351 | 76% | 34 | 71% |
|  | Passive | 53 | (9%) | 5 | 5% | 43 | 9% | 5 | 10% |
| Gender | Female | 339 | (56%) | 58 | 60% | 251 | 55% | 30 | 63% |
|  | Male | 265 | (44%) | 39 | 40% | 208 | 45% | 18 | 38% |
| Grade | First | 49 | (8%) | 17 | 18% | 30 | 7% | 2 | 4% |
|  | Second | 95 | (16%) | 18 | 19% | 73 | 16% | 4 | 8% |
|  | Third | 157 | (26%) | 19 | 20% | 121 | 26% | 17 | 35% |
|  | Forth | 155 | (26%) | 22 | 23% | 120 | 26% | 13 | 27% |
|  | Fifth | 93 | (15%) | 6 | 6% | 79 | 17% | 8 | 17% |
|  | Sixth | 55 | (9%) | 15 | 15% | 36 | 8% | 4 | 8% |
| BMI | Underweight or Normal weight | 399 | (66%) | 72 | 74% | 296 | 64% | 31 | 65% |
|  | Overweight | 141 | (23%) | 16 | 16% | 117 | 25% | 8 | 17% |
|  | Obese | 45 | (7%) | 7 | 7% | 30 | 7% | 8 | 17% |
|  | Severe Obesity | 8 | (1%) | 2 | 2% | 5 | 1% | 1 | 2% |
| Chronic Diseases | Bronchial asthma | 48 | (8%) | 8 | 8% | 37 | 8% | 3 | 6% |
|  | Chronic renal disease | 10 | (2%) | 5 | 5% | 4 | 1% | 1 | 2% |
|  | Diabetes Mellitus | 12 | (2%) | 3 | 3% | 8 | 2% | 1 | 2% |
|  | Heart disease | 12 | (2%) | 4 | 4% | 6 | 1% | 2 | 4% |
|  | Hypertension | 32 | (5%) | 8 | 8% | 23 | 5% | 1 | 2% |

N: Number, SD: Standard Deviation, BMI: Body Mass Index

### Distribution of parameters of the study sample according to state of positive PCR post COVID-19 vaccine

Table 2; shows that post vaccination breakthrough cases with positive PCR results were found in 74 (12%) participants after taking the 1st dose and in 49 (8%) after taking the 2nd dose, but as a predisposing factor; there was no statistically significant difference between the two groups in any of parameters. None had PCR testing after taking the 3^rd^ dose.

**Table 2:** Distribution of parameters of the study sample according to state of positive PCR post COVID-19 vaccine.

| Parameters |  | Positive PCR after 1st dose<br>N= 74 (12%) |  | Positive PCR after 2nd dose<br>N= 49 (8%) |  | Chi-square (p-value) |
| --- | --- | --- | --- | --- | --- | --- |
| **Gender** | Female | 36 | 49% | 26 | 53% | **0.22**<br>**(0.63)** |
|  | Male | 38 | 51% | 23 | 47% |  |
| **Grade** | First | 4 | 5% | 2 | 4% | **3.92**<br>**(0.41)** |
|  | Second | 17 | 23% | 8 | 16% |  |
|  | Third | 14 | 19% | 16 | 33% |  |
|  | Forth | 21 | 28% | 12 | 24% |  |
|  | Fifth | 10 | 14% | 4 | 8% |  |
|  | Sixth | 8 | 11% | 7 | 14% |  |
| **BMI** | Under or Normal weight | 48 | 65% | 30 | 61% | **1,868**<br>**(0.6)** |
|  | Overweight | 17 | 23% | 16 | 33% |  |
|  | Obese | 6 | 8% | 2 | 4% |  |
|  | Severe Obesity | 2 | 3% | 0 | 0% |  |
| **Chronic Disease** | Asthma | 8 | 11% | 6 | 12% | **1.7**<br>**(0.63)** |
|  | Chronic renal disease | 3 | 4% | 3 | 6% |  |
|  | Heart disease | 5 | 6% | 1 | 0% |  |
|  | Hypertension | 8 | 7% | 6 | 12% |  |
| **Smoking** | Current Smoker | 11 | 15% | 6 | 12% | **1.0**<br>**(0.57)** |
|  | Non smoker | 59 | 79% | 38 | 78% |  |
|  | Passive | 4 | 6% | 5 | 12% |  |

PCR: Polymerase Chain Reaction, N: Number, SD: Standard Deviation, BMI: Body Mass Index

### Distribution of number of symptoms according to number of received doses of COVID-19 vaccine

All participants were classified according to the number of symptoms they have developed post vaccination with different doses. The majority were symptomatic after taking the second dose but there was no statistically significant difference between these groups (P value =0.55), as shown in table 3.

**Table 3:** Distribution of number of symptoms according to number of received doses of COVID-19 vaccine.

| Number of Symptoms | One does | Two doses | Three doses | Total |
| --- | --- | --- | --- | --- |
| **0** | **0 (0%)** | **0 (0%)** | **0 (0%)** | **0 (0%)** |
| **1-2** | **55 (9%)** | **230 (38.1%)** | **20 (3.3%)** | **305 (50.4%)** |
| **3 or more** | **42 (7%)** | **229 (37.9%)** | **28 (4.7%)** | **299 (49.6%)** |
| **Total** | **97 (16%)** | **459 (76%)** | **48 (8%)** | **604** |

### Distribution of types of symptoms according to number of received doses of COVID-19 vaccine

The most frequent symptoms were fever, unusual fatigue, chills and persistent cough, as shown in table 4.

**Table 4:** Distribution of types of symptoms according to number of received doses of COVID-19 vaccine.

| Symptoms | one dose<br>N= 97 |  | Two doses<br>N= 459 |  | Three doses<br>N= 48 |  | All participants<br>N= 604 |  |
| --- | --- | --- | --- | --- | --- | --- | --- | --- |
| Persistent cough | 5 | 5% | 18 | 4% | 3 | 6% | 26 | 4.28% |
| Pins sensation | 0 | 0% | 4 | 1% | 0 | 0% | 4 | 0.66% |
| Depressed or hopeless | 2 | 2% | 2 | 0% | 0 | 0% | 4 | 0.66% |
| Diarrhoea | 2 | 2% | 2 | 0% | 0 | 0% | 4 | 0.66% |
| Chills | 6 | 6% | 21 | 5% | 2 | 4% | 29 | 4.77% |
| Feeling down | 2 | 2% | 3 | 1% | 0 | 0% | 5 | 0.82% |
| Loss of smell or taste | 1 | 1% | 7 | 0% | 1 | 2% | 9 | 1.48% |
| SOB | 1 | 1% | 8 | 2% | 0 | 0% | 9 | 1.48% |
| Unusual chest pain | 0 | 0% | 3 | 1% | 0 | 0% | 3 | 0.49% |
| Unusual fatigue | 8 | 8% | 84 | 18% | 4 | 8% | 96 | 15.79% |
| Unusual hair loss | 0 | 0% | 9 | 2% | 0 | 0% | 9 | 1.48% |
| Unusual muscle pains | 1 | 1% | 7 | 2% | 0 | 0% | 8 | 1.32% |
| Confusion | 0 | 0% | 1 | 0% | 1 | 2% | 2 | 0.33% |
| Earache | 1 | 1% | 1 | 0% | 0 | 0% | 2 | 0.33% |
| Fever | 63 | 65% | 275 | 60% | 34 | 71% | 372 | 61.18% |
| Headache | 0 | 0% | 7 | 2% | 1 | 2% | 8 | 1.32% |
| Lumps | 0 | 0% | 1 | 1% | 1 | 0% | 2 | 0.33% |
| Nausea or vomiting | 0 | 0% | 1 | 0% | 0 | 2% | 1 | 0.16% |
| Palpitations | 1 | 0% | 2 | 0% | 0 | 0% | 3 | 0.49% |
| Ringing in ears | 0 | 1% | 1 | 0% | 0 | 0% | 1 | 0.16% |
| Runny nose | 2 | 0% | 1 | 0% | 0 | 0% | 3 | 0.49% |
| Sore throat | 0 | 2% | 2 | 0% | 0 | 0% | 2 | 0.33% |
| Strange sensation in skin | 0 | 0% | 3 | 0% | 1 | 0% | 4 | 0.66% |
| hoarse voice | 2 | 0% | 0 | 1% | 0 | 2% | 2 | 0.33% |

## DISCUSSION

### Demographic& clinical data

In current study we aimed to assess the frequency and clinical characteristics of post COVID-19 vaccine breakthrough cases and to study any predictive risk factors. Out of total sample (604 participants), 97 (16%) showed symptoms of COVID-19 after taking the first dose of vaccine, while 459 (76%) after the second dose and only 48 (8%) after the third, this gives us a clue about the association between the possibility of showing symptoms and extended dose prospects. High percentage after taking the second dose of vaccine may be related to decrease commitment with precaution measures like wearing masks and social distancing in crowded places.

In current study; the majority were nonsmokers; this is due to the fact that the majority of our participants originally were nonsmokers. The mean age was 21.78 (± 3.26) which is close to what was found in another study in USA by (Feikin et. al, 2022) who found that younger people were more affected by such infections (29). Females were more than males which is agreed by a study done by Abou-Samra in Egypt in 2021 (30).

According to the BMI, 45 (7%) were obese while 399 (66%) were either underweight or normal. In a study done by (Gao M, et. al 2021); it was found that at a BMI of more than 23 kg/m^2^, there was a linear increase in risk of severe COVID-19 leading to admission to hospital and death, and a linear increase in admission to an ICU across the whole BMI range, which is not attributable to excess risks of related diseases (31).

Several studies have shown that obesity was emerging as a risk factor for susceptibility to COVID-19 (32-34). But a study done by (Petrilli et. al. 2020); showed that both underweight and obese patients with COVID-19 tend to develop acute lung injury compared with normal-weight patients. Underweight patients were more likely to develop a secondary infection than other patients (32).

In current study, bronchial asthma was the most frequent chronic condition found in 48(8%) and this may be related by some way to showing symptoms. Studies from Europe revealed contradictory data regarding asthma prevalence among COVID-19 patients with some yielding low percentages (<2%) by (Caminati in Italy 2020) (35), while in a study in Spain (Borobia et. al. 2020), found a higher prevalence (6%) (36).

More recent studies from the UK and the US revealed much higher rates of asthma among COVID-19 patients. An asthma prevalence of 14.4% was found in a US COVID-19 cohort by (Chhiba et. al. 2020) (37) exceeding the prevalence of the disease in the general population.

A similar percentage of 14% in asthma prevalence among COVID-19 patients was reported by (Docherty et. al. 2020) in the UK (38).

### Positive PCR Test

As we were trying to figure out the factors that contribute to develop breakthrough infection after vaccination, 74 (12%) of our participants showed positive PCR test after the first dose and 49 (9%) after the second dose, but there was no statistically significant association with any of the parameters as predictive risk factors (Age, Grade, Gender, BMI, Smoking and Chronic Diseases). In another study by (Jung J. 2021), 61% of the participants showed positive PCR after being fully vaccinated (39).

### Post Covid 19 Vaccine Symptoms

The majority of breakthrough cases were found among those who received two doses of COVID-19 vaccine and small numbers among those with three doses indicating the importance of booster doses of vaccine. In India, (Behera et. al) found that the majority of breakthrough cases was detected in fully vaccinated (40). The most frequent symptoms in this study were fever or feeling hot in 372 (61%) and unusual fatigue in 96 (15.6%) (table 4), but there was no statistically significant difference between number of vaccine doses and numbers of symptoms developed (p value =0.55) (table 3). In a study conducted by (Riad et. al, 2021), it was found that fatigue, headache and muscle pain were the most frequent symptoms (41).

## CONCLUSIONS

In current study; demographic factors showed no statistically significant impact on prevalence of post COVID-19 vaccination breakthrough cases. Despite this; number of participants who develop symptoms after the second dose of vaccine was high; and about 40% of them showed 3 or more symptoms. About half of participants showed symptoms even after being fully vaccinated.

## Data Availability

All data produced in the present work are contained in the manuscript

## RECOMMENDATIONS

1. Data in this study underscore the critical importance of continued public health mitigation measures (masking, physical distancing, daily symptom screening, and regular testing), even in environments with a high incidence of vaccination, until herd immunity is reached at large.
2. Further studies with larger sample size among health care workers are recommended.

